# Radiomics-Based Lung Nodule Classification with Stacking Ensembles

**DOI:** 10.1101/2025.04.28.25326620

**Authors:** Shao Ming Koh, Isaac Lam Hong Kei, Thanh Mai Nguyen

## Abstract

Radiomics, an emerging field in medical imaging, leverages advanced mathematical analysis to extract quantitative metrics from medical images, aiding in the early detection, diagnosis, and treatment of lung cancer. This study focuses on improving the risk prediction of small lung nodules using machine learning models. We employed a stacking ensemble approach, integrating Principal Component Analysis (PCA) for dimensionality reduction and selected features from the Small Nodule Radiomics-Predictive Vector (SN-RPV). Base models employed in the stacking ensemble were Support Vector Machine (SVM), Random Forest, k-Nearest Neighbors (KNN), and Naive Bayes classifiers. Despite the theoretical advantages of stacking ensembles, our models demonstrated poorer performance on the test set compared to the simpler SN-RPV model by Hunter et al. This outcome highlights the challenges of overfitting and underscores the importance of model simplicity and interpretability in clinical applications. Future research should explore alternative regularization techniques to improve the generalization of complex ensemble methods.

## Introduction

Radiomics, akin to genomics, proteomics, and metabolomics, is an emerging field in medicine that utilizes advanced mathematical analysis to provide a quantitative approach to medical imaging. By employing AI-based analysis techniques, radiomics extracts quantitative metrics and quantifies textural information from medical images [5]. This analysis can be performed across multiple imaging modalities, enabling an integrated approach that consolidates data from various imaging tools such as magnetic resonance imaging (MRI), computed tomography (CT), and positron emission tomography (PET). Traditionally, these imaging techniques are evaluated separately in conventional diagnostic imaging.

In the field of lung cancer, radiomics-based approaches are transforming disease management by improving early detection, diagnosis, prognosis, and treatment decision-making. One of the key clinical metrics for assessing cancer risk is the presence of lung nodules, with nodule size being a crucial determinant [4]. Nodule diameter is measured to classify nodules as either small or large.

The radiomics-based approach has emerged as an innovative method for improving lung cancer risk prediction. By extracting radiomic features from medical images and analyzing them using machine learning (ML) models, this approach aids in assessing the malignancy risk of indeterminate screen-detected pulmonary nodules [7]. Various ML models have been evaluated for this purpose, including eXtreme Gradient Boosted Trees, Random Forest, and Least Absolute Shrinkage and Selection Operator (LASSO) regression [6], as well as LASSO regression and K-Clustering [3].

Predicting malignancy in small nodules remains a significant challenge. Approximately 15% of individuals undergoing lung cancer screening require an early follow-up CT scan; however, the vast majority of these nodules ultimately prove to be benign. Conversely, some patients experience delayed lung cancer diagnoses and disease progression due to the growth of nodules initially classified as benign or indeterminate [1]. Despite the growing adoption of radiomics-based approaches, relatively few studies have focused specifically on small nodules.

Therefore, the objective of this project is to improve the risk prediction in small nodules of lung cancer using machine learning models via evaluating the performance of our proposed methodology against the benchmark model developed by Hunter et al. [3]. A distinguishing aspect of our approach is the incorporation of principal component analysis (PCA) for dimensionality reduction. PCA reduces computational complexity, mitigates overfitting, and enhances model performance by eliminating redundant features, while preserving the dataset’s essential structure. We assess the performance of four machine learning models within each scaling method: Support Vector Machine (SVM), Random Forest, k-Nearest Neighbors (KNN), and Naïve Bayes. Finally, we hypothesize that the introduction of a stacking ensemble will enhance predictive performance.

### Data Source

The dataset used in this study is publicly available and was originally collected and annotated by Hunter et al. [3]. The original dataset comprised CT images of lung nodules, which were processed to extract radiomic features using TexLab2.0. The final dataset includes 736 images, each containing 1998 radiomic features. These features provide quantitative information about the shape, texture, and intensity of the nodules. Out of the 736 images, 609 were allocated for training and internal validation, and the remaining 127 were set aside as a test set. Detailed information on the data collection, annotation processes, and radiomic feature extraction can be found in the original publication by Hunter et al. [3].

## Methodology

This project employs a stacking ensemble to enhance the classification of radiomics-based small lung nodules. The entire process is illlustrated in Figure 1.

**Figure 1:**
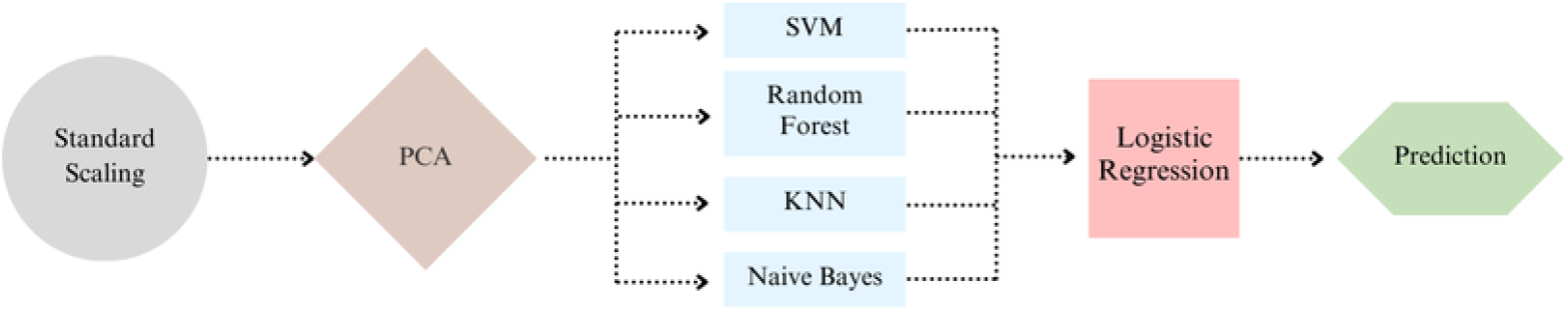
Flowchart of the stacking ensemble for small lung nodule classification

To ensure a robust and scalable workflow, machine learning models were constructed using Python’s Pipeline objects from the scikit-learn library. The data preprocessing involved standard scaling 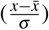, followed by Principal Component Analysis (PCA), before being fed into the base classifiers. The base classifiers included Support Vector Machine (SVM), Random Forest, K-Nearest Neighbor (KNN), and Naive Bayes.

Each pipeline is subsequently integrated into a hyperparameter optimization framework, RandomizedSearchCV, employing 5-fold cross-validation. This approach facilitates an efficient exploration of the hyperparameter space while minimizing the risk of overfitting. Each pipeline undergoes optimization for the number of components in the dimensionality reduction phase and model-specific parameters. The explored hyperparameter space for each base model is summarized in Table 1. The final predictions are generated by a Logistic Regression meta-model, which takes the probabilistic outputs of the base models as input. This is implemented using scikit-learn’s StackingClassifier.

**Table 1:** Hyperparameter space of base models.

| Model | Hyperparameter | Value |
| --- | --- | --- |
| SVM | C | uniform(0.1, 10) |
|  | Kernel | 'linear', 'rbf' |
| Random Forest | n_estimators | randint(50, 200) |
|  | max_depth | randint(3,20) |
| KNN | n_neighbors | randint(3, 20) |
|  | weights | 'uniform', 'distance' |
| Naive Bayes | NA | NA |

**Abbreviations:** SVM = Support Vector Machine; KNN = K-Nearest Neighbors; NA = Not Applicable

To further assess the effectiveness of an alternative dimensionality reduction method on this dataset, another stacking ensemble was developed using the same base classifiers, incorporating the five features identified by Hunter et al. [3] in the construction of his model. Additionally, the performance of the four base models was examined following preprocessing with spectral embeddings.

Receiver Operating Characteristic (ROC) curves were generated to assess the performance of all base models and the final stacking ensemble, with evaluation based on the Area Under the Curve (AUC). To estimate 95% confidence intervals, resampling was performed using 1000 bootstrap iterations. Threshold-based performance metrics including accuracy, precision, recall and F1-Scores, were reported using the ROC cutoff that maximized the Youden index on the training set. The Youden index is recognized as a balanced metric, providing equal weight to both sensitivity and specificity.

## Results

### Nodule Classification Statistics

Out of the 736 samples in the dataset, 377 nodules (51.2%) were identified as malignant, while 356 nodules (48.8%) were classified as benign. Within the training set of 609 samples, 305 (50.0%) were malignant and 304 (50.0%) were benign. In the test set, 72 (56.7%) nodules were malignant and 55 (43.3%) were benign. More detailed information on patient and nodule characteristics can be found in the paper by Hunter et al. [3].

### Base Model Results

Table 2 reports the training and cross-validation AUC-ROC scores of the base models employed in our stacking ensemble. The corresponding AUC-ROC plots are available in the supplementary materials (Figure S1 and Figure). Notably, validation scores in models pre-processed by spectral embedding declined significantly relative to their respective training scores, suggesting that overfitting has occurred. This is particularly evident in the spectral embedding-random forest (SE-RF) model, which achieved the highest training score (0.937) but a much lower validation score (0.644).

**Table 2:** Summary of base model training and cross-validation scores.

| Model | Train Score | Validation Score |
| --- | --- | --- |
| PCA-SVM | 0.843 | 0.816 |
| PCA-RF | 0.890 | 0.807 |
| PCA-KNN | 0.869 | 0.808 |
| PCA-NB | 0.834 | 0.790 |
| SE-SVM | 0.807 | 0.637 |
| SE-RF | 0.937 | 0.644 |
| SE-KNN | 0.894 | 0.649 |
| SE-NB | 0.797 | 0.640 |
| SNRPV-SVM | 0.850 | 0.724 |
| SNRPV-RF | 0.877 | 0.820 |
| SNRPV-KNN | 0.861 | 0.799 |
| SNRPV-NB | 0.848 | 0.744 |

**Abbreviations:** PCA = Principal Component Analysis; SE = Spectral Embedding; SNRPV = Selected variables that formulate Hunter et al.'s Small Nodule Radiomics Predictive Vector [3]; SVM = Support Vector Machine; RF = Random Forest; KNN = K-Nearest Neighbors; NB = Naive Bayes

### Stacking Ensemble

The stacking ensemble utilizing dimensionality reduction via PCA achieved an AUC-ROC score of 0.901 (95% CI: 0.876 to 0.924) on the training set and 0.774 (95% CI: 0.689 to 0.860) on the test set. The accuracy scores were 0.819 (95% CI: 0.788 to 0.849) for the training set and 0.685 (95% CI: 0.606 to 0.764) for the test set. In contrast, the stacking ensemble incorporating selected variables from the SN-RPV model attained an AUC-ROC score of 0.894 (95% CI: 0.869 to 0.920) on the training set and 0.769 (95% CI: 0.682 to 0.848) on the test set. The accuracy scores for this model were 0.816 (95% CI: 0.785 to 0.849) on the training set and 0.717 (95% CI: 0.638 to 0.795) on the test set. The precision, recall, and F1 scores are detailed in Table 3.

**Table 3:** Classification performance metrics of the stacking ensesmbles on training and test sets.

| Model | Classification | Precision (train, test) | Recall (train, test) | F1-Score (train, test) |
| --- | --- | --- | --- | --- |
| PCA | Benign | 0.84, 0.63 | 0.79, 0.65 | 0.81, 0.64 |
|  | Malignant | 0.80, 0.73 | 0.85, 0.71 | 0.82, 0.72 |
|  | Macro average | 0.82, 0.68 | 0.82, 0.68 | 0.82, 0.68 |
| Stacked SN-RPV | Benign | 0.87, 0.70 | 0.74, 0.60 | 0.80, 0.65 |
|  | Malignant | 0.78, 0.72 | 0.89, 0.81 | 0.83, 0.76 |
|  | Macro average | 0.82, 0.61 | 0.82, 0.70 | 0.82, 0.71 |

**Abbreviations:** PCA = Stacking ensemble utilizing Principal Component Analysis; Stacked SN-RPV = Stacking ensemble incorporating selected features from Hunter et al.'s SN-RPV [3]

### Baseline Comparison

The baseline performance of Hunter et al.’s small nodule radiomics-predictive-vector (SN-RPV) [3] is provided in the supplementary materials (Table S1). In comparison, our stacking ensembles showed superior performance across all macro average metrics on the training set. However, they exhibited reduced performance on the test set. Similarly, the AUC-ROC and accuracy scores of our stacking ensembles were greater than those of the baseline model on the training set (Figure 2a), but slightly weaker on the test set (Figure 2b).

**Figure 2:**
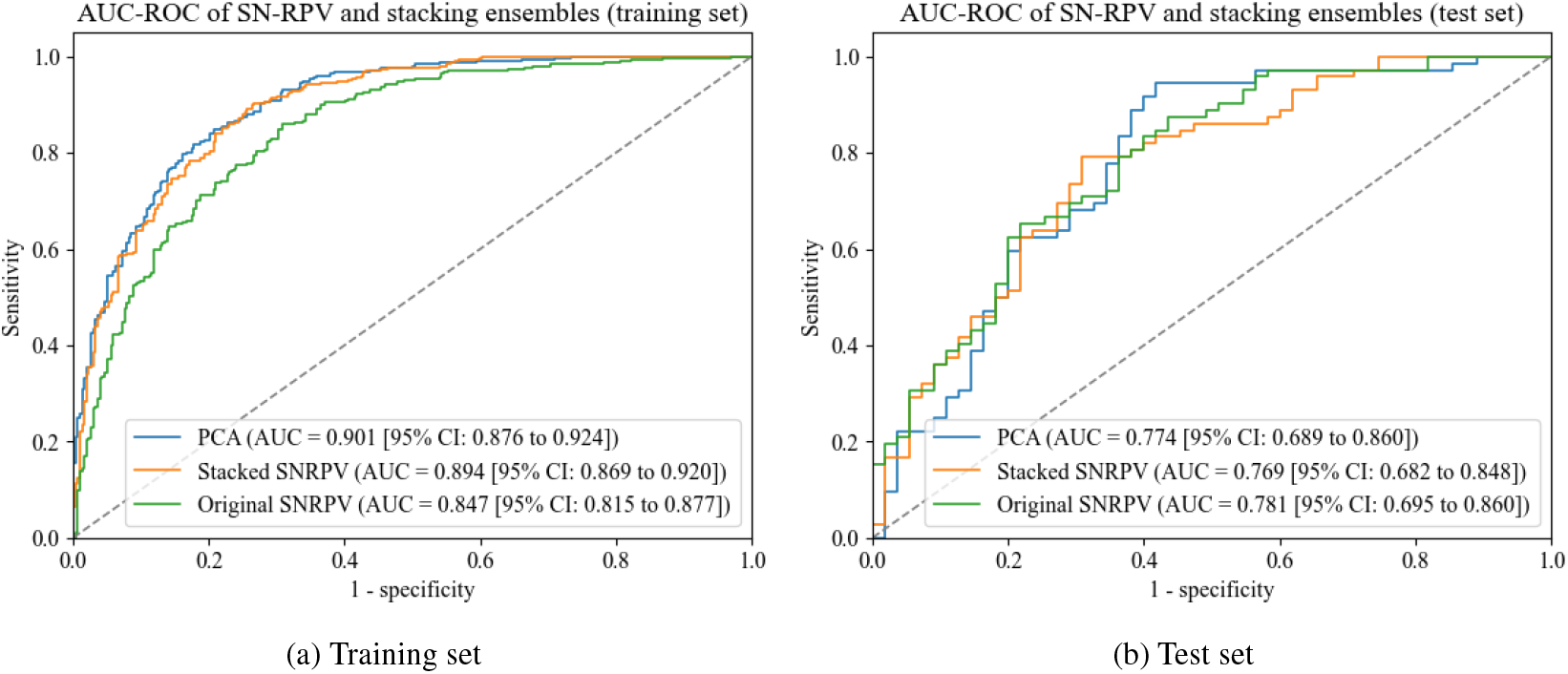
AUC-ROC plot of stacking ensemble on the (a) training set, (b) test set **Abbreviations:** AUC = Area Under Curve; CI = Confidence Interval; PCA = Principal Component Analysis; Original SNRPV = Hunter et al.’s [3] original model, Small Nodule Radiomics-Predictive Vector (SN-RPV); Stacked SNRPV = The stacking ensemble taking variables from the SN-RPV as input

## Discussion

Overall, the results of the stacking ensembles were surprising. Despite our initial expectations that they would outperform Hunter et al.’s SN-RPV [3], our models demonstrated relatively poorer performance on the test set. This outcome was unexpected, given that stacking ensembles are typically known for their robust predictive performance by leveraging the strengths of individual models. Stacking enembles are capable of achieving high predictive performance because different models may excel on various subsets of the data. By combining these models, stacking can handle diverse data more effectively, thereby enhancing generalization. Additionally, aggregating predictions from multiple models helps reduce the variance of the final prediction, leading to more stable and reliable results [2]. Thus, we had carefully selected diverse base models according to their distinct algorithmic approaches, aiming to leverage their complementary strengths.

Despite demonstrating strong performance on the training set, the stacking ensembles exhibited poor performance on the test set, indicating potential overfitting to the training data. This was evident from the significant drop in accuracy and AUC-ROC scores when comparing results from the training and test set. While stacking ensembles are designed to enhance generalization and mitigate overfitting by leveraging the predictions of diverse models, the increased complexity introduced by multiple models can inadvertently capture noise, leading to overfitting. Additionally, the relatively small training set, comprising 609 examples, may not provide sufficient data to learn generalizable patterns, further increasing the risk of overfitting.

Our ensemble model utilizing PCA for dimensionality reduction did not significantly outperform the ensemble model incorporating the selected SN-RPV variables, and it also failed to surpass hunter et al.’s SN-RPV [3]. PCA, unlike the feature selection method employed by Hunte et al. [3], preserves the directions that capture the most variance in the data [2]. Our intention was to minimize information loss during dimensionality reduction, ensuring that the essential structure and key characteristics of the data were maintained. However, compared to the feature selection approach, PCA lowers model interpretability since the principal components are combinations of the original features. This makes it challenging to understand the contribution of individual features to the model’s predictions. PCA is based on the assumption that the key characteristics of the dataset lie along the axes of greatest variance. However, this assumption may not necessarily hold for all datasets. We also explored spectral embeddings for dimensionality reduction, but they performed worse on the base models, leading us to exclude them from the stacking ensemble.

Despite the added complexity of our stacking ensembles, they have failed to improve radiomics-based classification of small lung nodules as compared to the SN-RPV [3]. This makes the SN-RPV the preferred model, not only for its predictive performance but also its simplicity, and interpretability. The practical implications of our findings suggest that while stacking ensembles have theoretical advantages, their application in radiomics-based lung nodule classification may require larger datasets and more refined tuning to avoid overfitting. This highlights the importance of simplicity and interpretability in clinical models, as demonstrated by the superior performance of the SN-RPV model [3].

## Conclusion

This study investigated the use of stacking ensembles for radiomics-based lung nodule classification. Despite their theoretical advantages, our stacking models underperformed compared to the simpler SN-RPV model by Hunter et al. [3]. We also explored PCA for dimensionality reduction, but it did not significantly improve performance. These results highlight the challenges of overfitting and the importance of model simplicity and interpretability in clinical applications. Future research should focus on alternative regularization techniques to improve the generalization of complex ensemble methods. Ultimately, our findings suggest that simpler models like SN-RPV can be more effective and reliable in certain contexts.

## Data Availability

The radiomics data utilized in this study were initially deposited by Hunter et al. [3] in the Mendeley database, accessible via the accession code 10.17632/rxn95mp24d.1. Additionally, the Python scripts employed for model development can be found at https://github.com/isaaclhk/Projects/tree/main/lungnoduleclassification.

## Supplementary Materials

**Figure S1:**
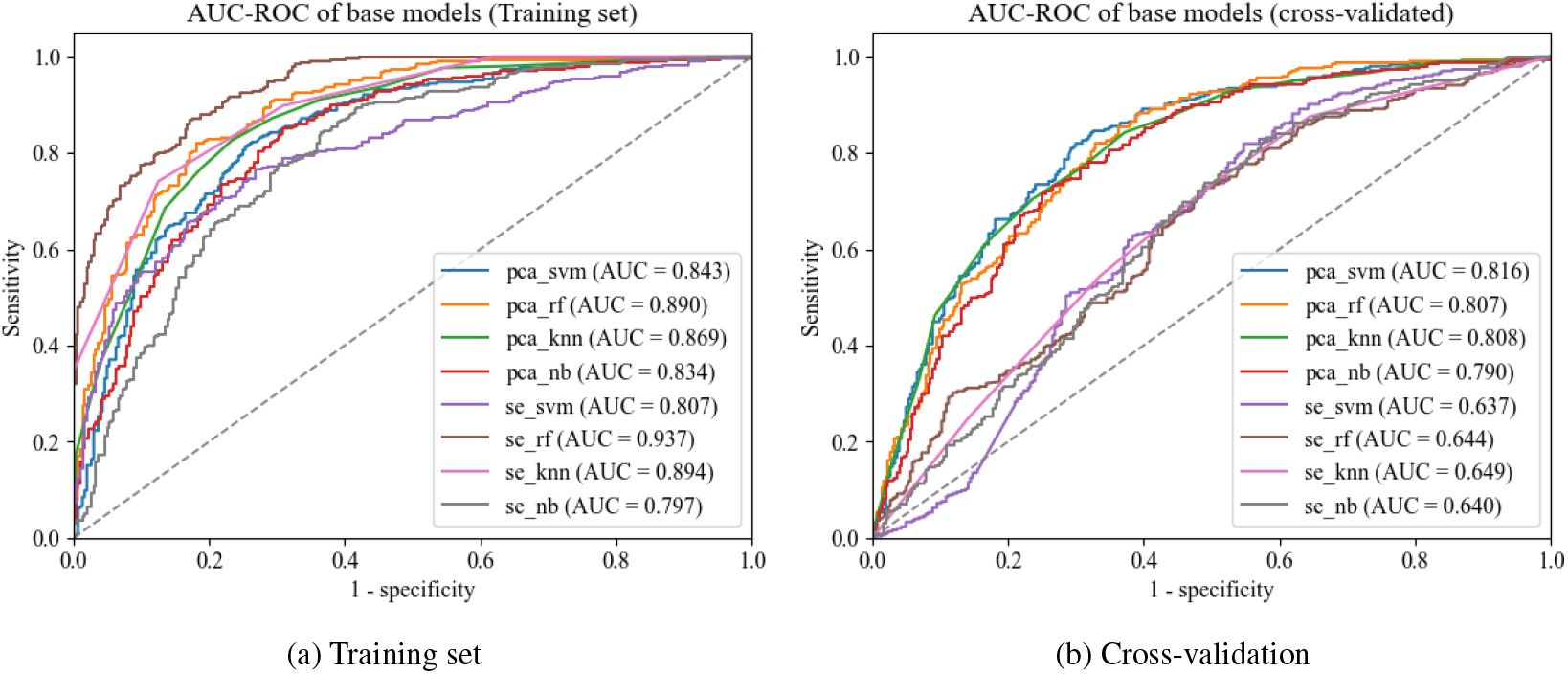
AUC-ROC of base models on the (a) training set and (b) cross-validation data after preprocessing with PCA or SE **Abbreviations:** AUC-ROC = Area Under the Curve of the Receiver Operating Characteristic; knn = K-Nearest Neighbor; nb = Naive Bayes; pca = Principal Component Analysis; rf = Random Forest; se = Spectral Embedding; svm = Support Vector Machine

**Figure S2:**
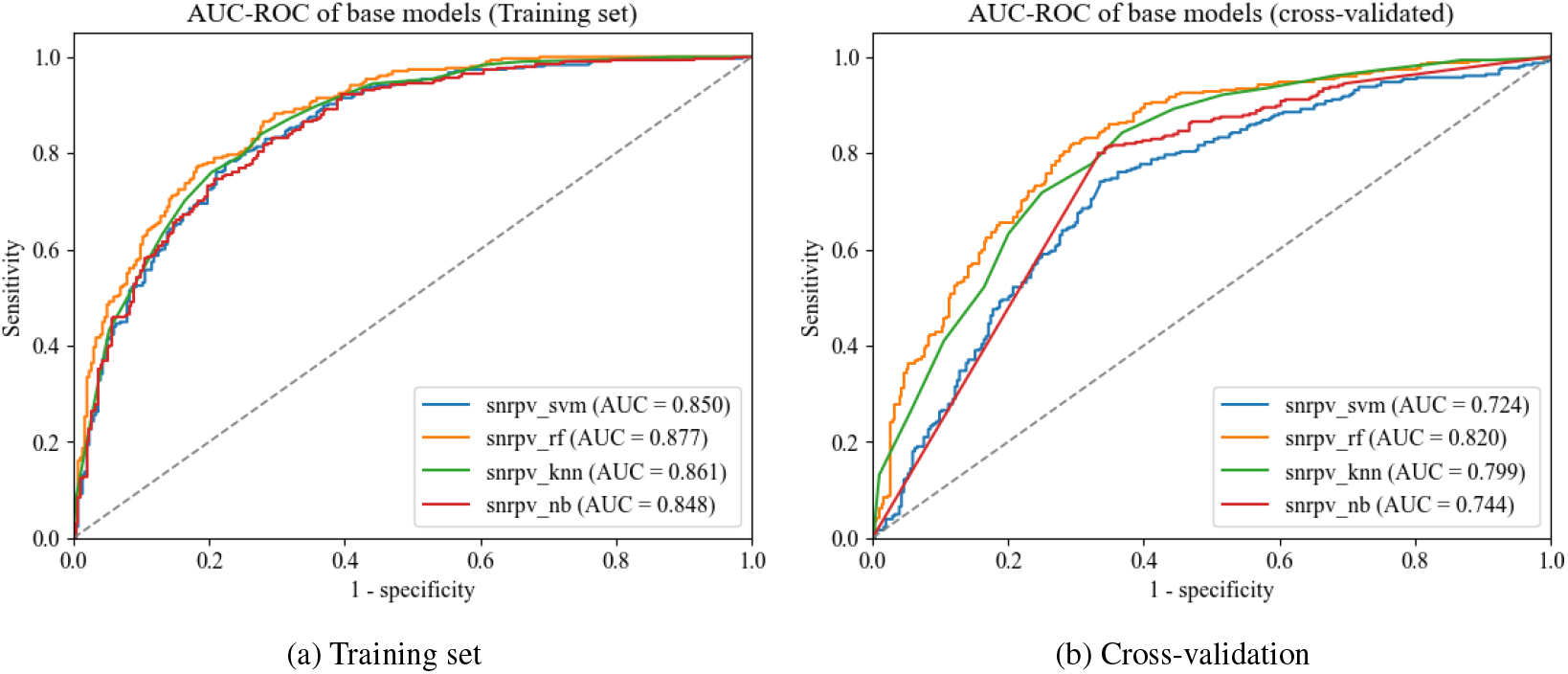
AUC-ROC of base models on the (a) training set and (b) cross-validation data taking selected variables from the SN-RPV as input **Abbreviations:** AUC-ROC = Area Under the Curve of the Receiver Operating Characteristic; knn = K-Nearest Neighbor; nb = Naive Bayes; rf = Random Forest, snrpv = Small Nodule Radiomics-Predictive-Vector; svm = Support Vector Machine

**Table S1:** Classification performance metrics of SN-RPV on training and test sets.

| **Classification** | **Precision (train, test)** | **Recall (train, test)** | **F1-Score (train, test)** |
| --- | --- | --- | --- |
| Benign | 0.83, 0.76 | 0.69, 0.56 | 0.75, 0.65 |
| Malignant | 0.74, 0.72 | 0.86, 0.86 | 0.79, 0.78 |
| Macro average | 0.78, 0.74 | 0.77, 0.71 | 0.77, 0.72 |

